# Convalescent Plasma to Treat COVID-19: Chinese Strategy and Experiences

**DOI:** 10.1101/2020.04.07.20056440

**Authors:** Shiyao Pei, Xi Yuan, Zhimin Zhang, Run Yao, Yubin Xie, Minxue Shen, Bijuan Li, Xiang Chen, Mingzhu Yin

## Abstract

The COVID-19 is currently spreading around the world, which has posed significant threats to global health and economy. Convalescent plasma is confirmed effective against the novel corona virus in preliminary studies. In this paper, we first described the therapeutic schedule, antibody detection method, indications, contraindications of the convalescent plasmas, and reported the operability of the treatment by case study.

## The body of the text

In December 2019, pneumonia associated with a novel corona virus 2019 (COVID-19) caused an outbreak, which has posed significant threats to global health and economy^1^. As of 6th April 2020, according to the World Health Organization Situation Report, this epidemic had spread to more than 180 countries with 1,113,758 confirmed cases, including 62,784 deaths^2^. The reported severely/critically ill case ratio is approximately 7-10%^3^ and mainly rely on the supportive care since specific drugs of COVID-19 are still being researched. On March 4, 2020, in order to improve the therapeutic effect of COVID-19, the National Health Commission of the People’s Republic of China organized Chinese experts to make revisions of the “Clinical treatment of COVID-19 Convalescent Plasma (the second trial edition)”^4^. On March 24, 2020, FDA approved the testing of convalescent plasmas for patients with serious or immediately life-threatening COVID-19 infections^5^. To date, thousands of convalescent plasmas have been collected and remarkable efficacy have been achieved in severely and critically ill COVID-19 patients in China. To share Chinese experience with the world and to standardize the treatment, we summarized the therapeutic schedule of the convalescent plasmas as follows (Fig.1).

**Fig. 1.**
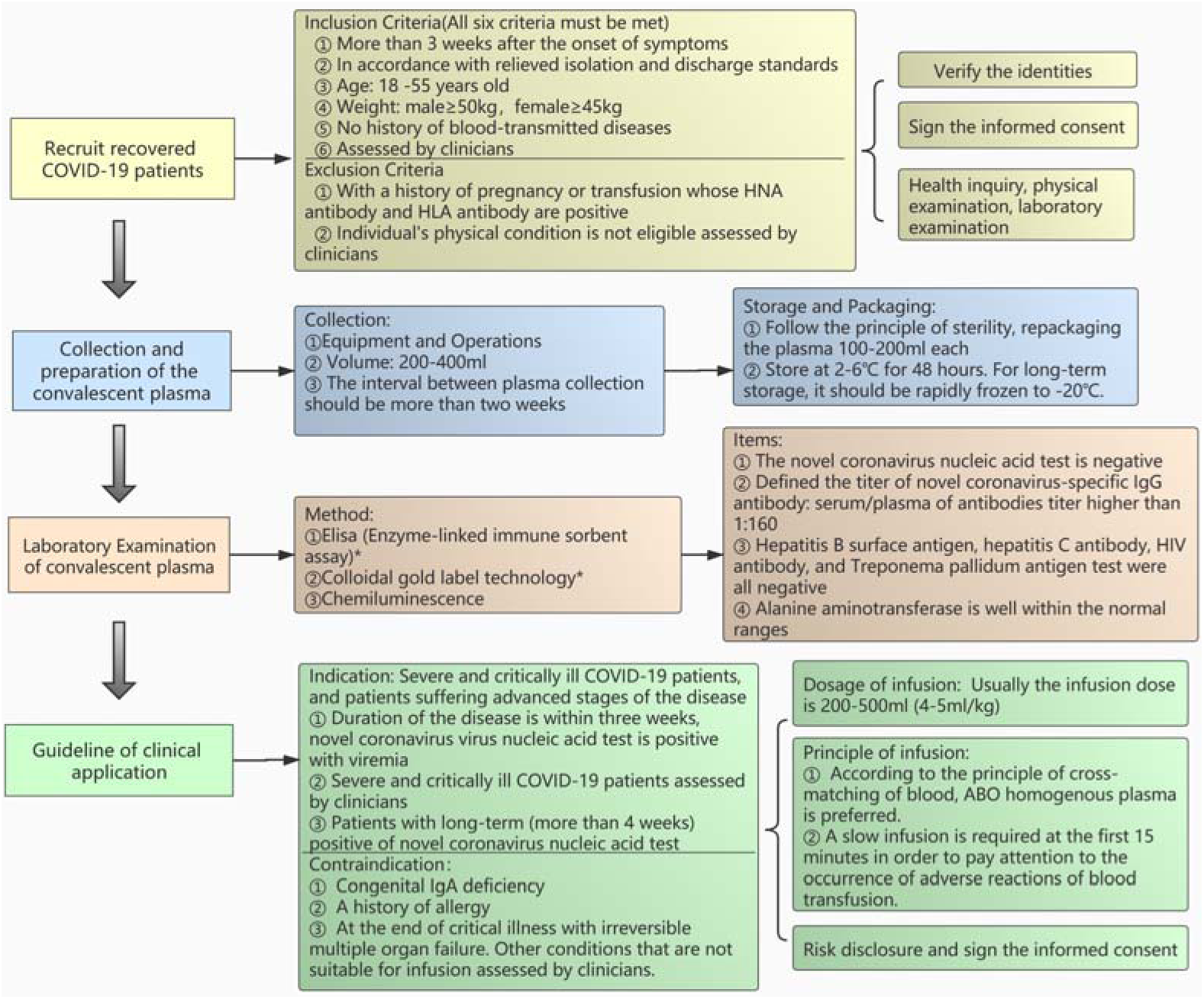
The standardized flow chart of the Convalescent Plasma transfusion.

## 1. Recruit recovered COVID-19 patients

### 1.1 Inclusion Criteria (All six criteria must be met)

□ More than 3 weeks after the onset of symptoms of the COVID-19 and complete resolution of symptoms at least 14 days prior to donation.
□ In accordance with relieved isolation and discharge standards following the latest version of the therapeutic schedule.
□ Age: 18-55 years old.
□ Weight: male≥50kg, female≥45kg.
□ No history of blood-transmitted diseases.
□ Eligible donors must be assessed by clinicians according to treatment.

### 1.2 Exclusion Criteria

□ With a history of pregnancy or transfusion whose HNA antibody and HLA antibody are positive.
□ Individual’s physical condition is not eligible assessed by clinicians.

### 1.3 Verify the identities

### 1.4 Sign the informed consent

### 1.5 Health inquiry, physical examination, laboratory examination of blood samples (refer to technical operation procedures of blood station)

## 2. Collection and preparation of the convalescent plasma

### 2.1 Collection

□ Equipment and Operations: fully automatic apheresis machine or a fully automatic blood cell separator (refer to technical operation procedures of blood station).
□ Volume: 200-400ml (The exact volume should be assessed by clinicians).
□ The interval between plasma collection should be more than two weeks.

### 2.2 Storage

□ Follow the principle of sterility, repackaging the plasma 100-200ml each.
□ Store at 2-6□ for 48 hours. For long-term storage, it should be rapidly frozen to −20□.

### 2.3 Packaging

Labelling Requirements: refer to technical operation procedures of blood station.

## 3. Laboratory Examination of convalescent plasma

### 3.1 Method

Elisa (Enzyme-linked immune sorbent assay)*, Colloidal gold label technology*, Chemiluminescence.

*Note: We recommend using ELISA to detect the novel coronavirus antibody titer since the colloidal gold method is not suitable for titer detection and the false negative rate is high. The analysis of 17 samples showed that the positive rate and sensitivity of ELISA were significantly better than colloidal gold (The specific data is shown in Supplementary Table 1).

### 3.2 Items

□ The novel coronavirus nucleic acid test is negative.
□ Defined the titer of novel coronavirus-specific IgG antibody: serum/plasma of antibodies titer higher than 1:160.
□ Hepatitis B surface antigen, hepatitis C antibody, HIV antibody, and Treponema pallidum antigen test were all negative.
□ Alanine aminotransferase is well within the normal ranges.

## 4. Guideline of clinical application

### 4.1 Indication

Severe and critically COVID-19 patients, and patients suffering advanced stages of the disease.

□ Duration of the disease is within three weeks, novel coronavirus virus nucleic acid test is positive with viremia.
□ Severely and critically ill COVID-19 patients assessed by clinicians.
□ Patients with long-term (more than 4 weeks) positive of novel coronavirus nucleic acid test (for details please refer to patient 2 in Fig.2).

**Figure 2.**
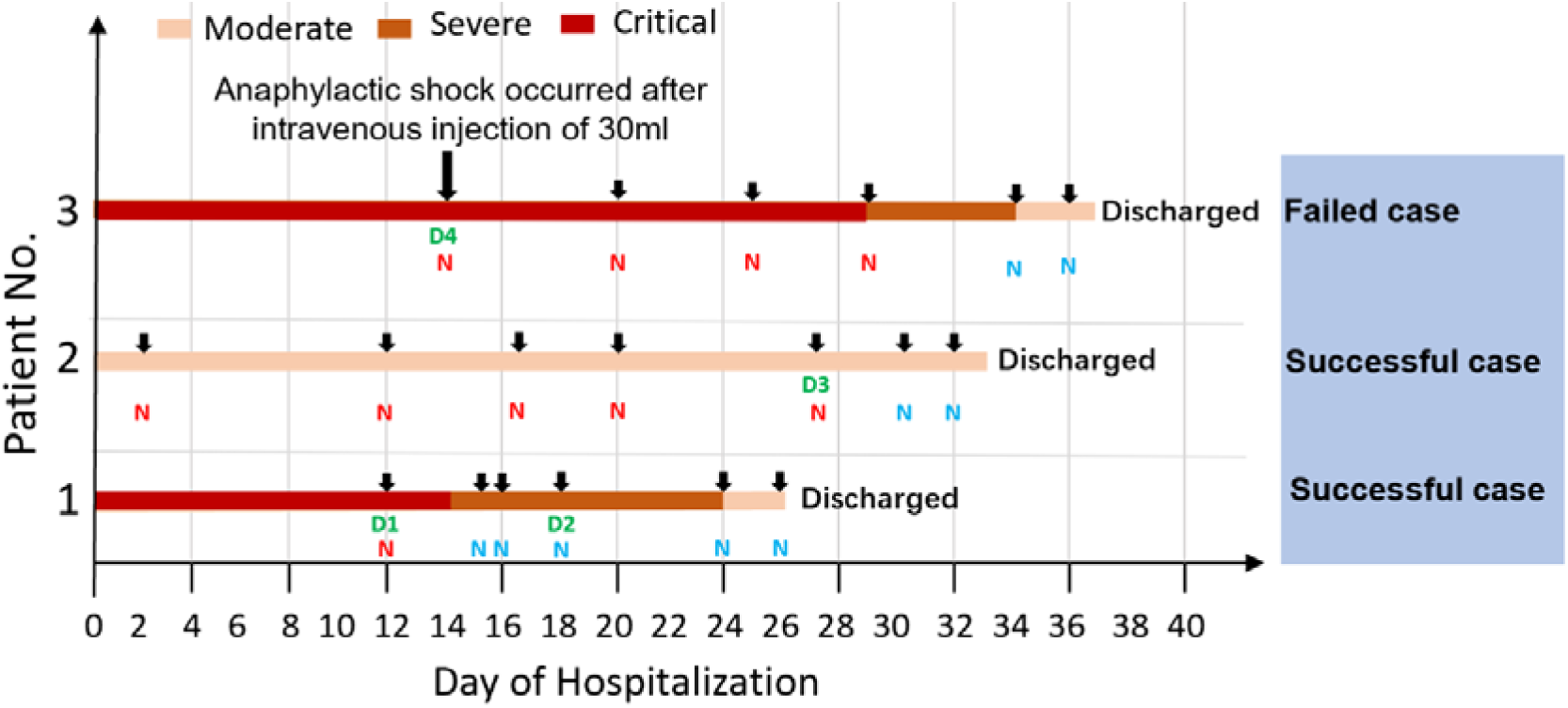
Outcomes for individual patients included in three cases. Donor and receiver detail information see table 1; N, Nucleic acid test positive; N, Nucleic acid test negative; D, Donor patient

### 4.2 Contraindication

□ Congenital IgA deficiency.
□ A history of allergy including plasma infusion, human plasma protein products, sodium citrate. Plasma inactivated by methylene blue virus is strictly prohibited in patients with methylene blue allergy. Other history of severe allergies and contraindications.
□ At the end of critical illness with irreversible multiple organ failure. Other conditions that are not suitable for infusion assessed by clinicians.

### 4.3 Dosage of infusion

According to the clinical status and the patient’s weight. Usually the infusion dose is 200-500ml (4-5ml/kg).

### 4.4 Principle of infusion

□ According to the principle of cross-matching of blood, ABO homogenous plasma is preferred.
□ A slow infusion is required at the first 15 minutes in order to pay attention to the occurrence of adverse reactions of blood transfusion.

### 4.5 Risk disclosure and sign the informed consent

## 5. Three clinical cases of the convalescent plasma transfusion

Here we show the outcomes of three individual patients with convalescent plasma transfusion (Fig.2), and the characteristics of the donor/receiver are shown in the supplementary table 2 (The participants gave their written informed consent and approved by the hospital ethics committee). The first patient was treated by convalescent plasma on the 12^th^ day of admission, the viral nucleic acid test became negative and the critical illness turned to severe. After the second therapy was conducted, the patient had improved significantly and the discharge criteria were met on 26th day. We can learn from this case that convalescent plasma treatment can quickly turn the viral nucleic acid test negative and significantly reduced symptoms. The second case was a patient whose nucleic acid test was consistently positive with moderate symptoms, after treatment on 27^th^ day, the viral nucleic acid test turned negative four days later. Therefore, in order to save medical resources and reduce the transmission of infection, it is advisable to use convalescent plasma treatment on patients with long-term (more than 4 weeks) positive nucleic acid test of novel coronavirus. In the third case, the plasma donor was a 51-year-old woman with a history of pregnancy (supplementary table 2), the receiver had a severe anaphylactic shock after intravenous injection of 30ml convalescent plasma. This empirical case warned us that the selection of plasma donors should be strictly in accordance with the inclusion criteria.

The use of convalescent plasma has a long history, at end of the 19th century, researcher found that recovery patients’ plasma was effective in diphtheria and tetanus patients. Use of convalescent plasma has been studied in outbreaks of other respiratory infections, including the 2003 SARS-CoV-1, 2009-2010 H1N1 influenza virus pandemic, and the 2012 MERS-CoV epidemic^6^. Previous studies showed a shorter hospital stay and lower mortality with no adverse events or complications in patients treated with convalescent plasma treatment than those who were not^7^. Additionally, viral load after convalescent plasma treatment was significantly lower on days 3, 5, and 7 after intensive care unit admission^8^. What’s more, we found that patients with long-term positive nucleic acid test of novel coronavirus turn negative earlier after convalescent plasma treatment than those without convalescent plasma treatment. Furthermore, asymptomatic patients with hypoimmunity, such as the elderly, children and patients with underlying diseases such as diabetes, hepatitis, AIDS, heart disease, tuberculosis, malignant tumor, etc., preferred to use convalescent plasma once the nucleic acid test is positive. In the fighting against emerging and pandemic COVID-19, the advantages of the convalescent plasma have been validated by the practice and clinical outcome in China. Therefore, it provides an unprecedented opportunity to perform clinical studies and trials of the efficacy to the convalescent plasma during the pandemic of COVID-19, meanwhile, it is worthwhile to utilize the safety and efficacy of convalescent plasma transfusion in SARS-CoV-2-infected patients globally.

## Data Availability

All the data referred in the manuscript is available

## Authors and contributions

Xi Yuan, Zhimin Zhang collected the data. Shiyao Pei and Mingzhu Yin drafted the manuscript. Run Yao, Yubin Xie, Minxue Shen contributed to the interpretation of the results and critical revision of the manuscript for important intellectual content and approved the final version of the manuscript. Bijuan Li, Xiang Chen were involved in data cleaning, and verification. All authors have read and approved the final manuscript.

## Funding

No.

## Declaration of interests

All authors declare no competing interests.

## Acknowledgements

We thank all the hospital staff members for their efforts in collecting the information that used in this study; thank the patients who participated in this study, their families, and the medical, nursing, and research staff at the study centers.

## Data sharing

No additional data are available.

## Notes

### Competing Interest Statement

The authors have declared no competing interest.

### Funding Statement

No funding

